# The KIND cohort profile: longitudinal assessment of glycaemic management and neurophysiological outcomes in paediatric type 1 diabetes in Switzerland

**DOI:** 10.64898/2026.01.11.26343881

**Authors:** Marc Robin Gruener, Erin A. West, Ute Muhitira, Katrin Heldt, Sarah S. Oberhauser, Dagmar l’Allemand-Jander, Philip J. Broser, the KIND Study Group

**Affiliations:** Children’s Hospital of Eastern Switzerland, St. Gallen, Switzerland; Clinical Trials Unit, HOCH Health Ostschweiz, St. Gallen, Switzerland; Metabolic Centre St. Gallen, Friendlydocs AG, St. Gallen, Switzerland; Medical Faculty, University of Basel, Basel, Switzerland

**Keywords:** Pediatrics, Diabetes Mellitus, Type 1, Diabetic Neuropathies, Nerve Conduction Studies, Peripheral Nervous System

## Abstract

1

**Purpose:** The KIND (KINder mit Diabetes) cohort investigates diabetic peripheral neuropathy (DPN) in paediatric type 1 diabetes (T1D). Current guidelines recommend DPN screening at puberty or from 11 years and 2-5 years after T1D diagnosis, yet subclinical neurophysiological changes occur within the first 2 years. The cohort examines: (1) longitudinal associations between glycaemic metrics (HbA1c and continuous glucose monitoring-derived variability metrics) and peripheral nerve function and structure; (2) comparative predictive value of different variability metrics; (3) developmental trajectories of nerve maturation in T1D versus controls; (4) effects of residual beta-cell function on neuropathy progression; and (5) how early signs of DPN differ between patients with multiple daily injections (MDI) and continuous subcutaneous insulin infusion (CSII) therapy.

**Participants:** This prospective cohort, initiated in June 2019, continuously enrols children and young adults with T1D (≤21 years) at two Swiss centres. The care-embedded design integrates research into quarterly diabetes care and annual comprehensive assessments. So far, 141 T1D cohort participants (median age: 12.2; IQR: [8.4; 14.3] years; 47.5 % female) and 103 healthy controls (10.9 [7.5; 14.2] years; 53.4 % female) were recruited. Controls for neurophysiological examinations comprise measurements from the healthy, contralateral side of children with limb injuries in the surgical outpatient clinic of the Children’s hospital of Eastern Switzerland (OKS). Multimodal assessments comprise nerve conduction studies (peroneal, tibial, median motor and sensory) and high-resolution ultrasound, with development-adjusted analyses distinguishing diabetes effects from normal growth.

**Findings to date:** Cross-sectional analysis showed reduced nerve conduction velocities across all nerves, particularly peroneal in T1D patients (n=53) compared to healthy controls (n=50). Height-adjusted peroneal velocity (dNCV) correlated negatively with glucose variability (SD: r=-0.45, p=0.009), HbA1c (r=-0.27, p=0.049). During the first five years, dNCV correlated negatively with diabetes duration (r=-0.41, p=0.004), independent of glycaemic control. The cross-sectional area of the median nerve increased on average by 0.217 mm² per 1% HbA1c (p=0.004) and was already detectable with diabetes duration <2 years. Longitudinal analysis (n=45, 21.4±8.6 months) demonstrated that HbA1c changes predicted dNCV changes (β=-0.59, p=0.014), and in some patients, early impairment was reversible with improved glycaemic control.

**Future plans:** A planned 2026 extension of this continuously recruited and prospectively followed cohort will integrate physical activity measures (Swiss National Science Foundation Grant 10006264). Future analyses will compare glycaemic variability metrics as predictors of functional and structural nerve changes, investigate their temporal relationships as well as influencing factors, and examine residual beta-cell function effects across developmental stages.

All data produced in the present study may be made available upon reasonable request and in accordance with legal and ethical requirements.

## 2 STRENGTHS AND LIMITATIONS OF THIS STUDY

### Strengths

- Longitudinal clinical and neurophysiological investigation from type 1 diabetes diagnosis through adolescence captures peripheral neuropathy development compared with age-matched healthy controls.
- Integration of nerve conduction studies with high-resolution ultrasound provides comprehensive functional and structural assessment using non-invasive and development-adjusted analyses.
- Continuous glucose monitoring and quarterly HbA1c enable precise quantification of glycaemic control and comparison of multiple variability metrics as neuropathy predictors.

### Limitations

- Two-centre regional study limits geographic and ethnic diversity.
- A broad variety of continuous glucose monitoring devices across manufacturers complicate metric standardization despite harmonization efforts.

## 3 INTRODUCTION

Diabetic peripheral neuropathy (DPN) is the most common microvascular complication of diabetes, affecting nearly half of all patients during their lifetime.^1–4^ In children and adolescents with type 1 diabetes (T1D), DPN manifests as dysfunction of peripheral nerves, typically in a “stocking-and-glove” distribution, and is associated with impaired balance, sensory loss, and pain.^5^ Prevalence rates in paediatric populations range from 7 % to over 57%, depending on diagnostic criteria and patient characteristics.^6^ Longitudinal studies demonstrate that DPN begins early and progresses throughout childhood: Hyllienmark et al.^7^ reported a 15 % incidence of clinical neuropathy within 13 years, while Hajas et al.^8^ observed subclinical DPN prevalence increasing from 24 to 63 % over ten years. Despite this early onset and progressive nature, current International Society for Pediatric and Adolescent Diabetes (ISPAD) guidelines recommend screening at puberty or from 11 years of age and two to five years post-diagnosis of T1D. However, recent evidence demonstrates that subclinical changes are detectable within the first two years following diagnosis of T1D^7,9,10^, highlighting the need for early detection methods and better understanding of modifiable risk factors before irreversible nerve damage occurs.

Several fundamental questions remain unanswered regarding DPN pathogenesis and progression in children and adolescents. First, while HbA1c predicts long-term complication risk^11,12^, glycaemic variability (GV) has emerged as a potentially independent contributor to neuronal injury.^9^ Rapid glucose oscillations, common in paediatric T1D, may induce oxidative stress and pro-inflammatory signalling^13–15^, but it remains unclear how well GV metrics (standard deviation, coefficient of variation, time-in-range, or more complex indices^16–19^) predict neurophysiological changes over time. The widespread use of continuous glucose monitoring (CGM) enables detailed GV assessment, yet device heterogeneity across manufacturers complicates metric standardization and comparability^20^. Second, distinguishing diabetes-related nerve impairment from normal developmental changes poses a unique paediatric challenge. Nerve conduction velocity (NCV) increases dramatically from 20 m/s at birth to 45 m/s by age two, with maturation continuing into puberty, while nerve cross-sectional area expands proportionally with body growth.^21^ Whether age at diabetes diagnosis influences neuropathy trajectories through interactions with nervous system development remains unknown. Third, residual beta-cell function during the remission phase may be protective against complications^22^, and preliminary observations suggest that peroneal height-adjusted NCV (dNCV; defined as the difference between measured and, given a patient’s height, expected NCV) declines independently of glycaemic control during remission phase cessation.^9^ Whether C-peptide levels modify neuropathy progression requires longitudinal investigation. Finally, standard clinical screening tools (monofilament testing, vibration perception)^23^ have overall poor sensitivity in children^24,25^, while more sensitive functional (nerve conduction studies; NCS) and structural (high-resolution ultrasound) assessments can detect subclinical changes^9,24,25^, but how these modalities relate to each other temporally remains poorly characterized.

Addressing these knowledge gaps requires a cohort design that captures both disease progression and developmental maturation over extended periods. The KIND cohort employs continuous enrolment from T1D diagnosis onward, enabling investigation of neuropathy from its earliest stages through adolescence and young adulthood. This approach spans the full spectrum of diabetes duration and developmental stages. Integration into routine clinical care, quarterly visits with annual comprehensive assessments, maximizes retention and minimizes patient and family burden.

A control group of children undergoing neurophysiological evaluation for unilateral fractures provides normative developmental reference values, taken from the unaffected limb, for these adjustments. This comprehensive, care-embedded approach enables investigation of how glycaemic patterns interact with developmental stage and residual beta-cell function to influence peripheral nerve structure and function.

The KIND cohort investigates five interrelated objectives: first, to characterize longitudinal associations between glycaemic control metrics (HbA1c and GV indices) and peripheral nerve function and structure in children and adolescents with T1D; second, to compare the predictive value of different GV metrics for neurophysiological outcomes; third, to assess the influence of residual beta-cell function (C-peptide) on neuropathy progression, particularly during the remission phase; and fourth, to delineate developmental trajectories of peripheral nerve maturation in T1D compared with controls, accounting for age, pubertal stage, and diabetes duration. Fifth, the study examines how glycaemic control and early signs of neuropathy differ between MDI and CSII therapy. By evaluating these objectives, the cohort is expected to identify modifiable risk factors for early intervention and provide information for optimising strategies of paediatric diabetes treatment.

## 4 COHORT DESCRIPTION

### 4.1 Study Design and Setting

The KIND longitudinal cohort was started in June 2019. This ongoing prospective observational cohort study is conducted at two centres in Eastern Switzerland: The Children’s Hospital of Eastern Switzerland (OKS) in St. Gallen and the St. Gallen Metabolic Centre (SGMC).

The cohort is designed as a “care-embedded” study, integrating research data collection into the routine clinical care pathway for paediatric patients with T1D. This approach maximizes participant retention while minimizing burden on families.

Both centres follow the most current ISPAD guidelines 2022/2024 for diabetes management. T1D diagnosis is confirmed according to ISPAD clinical and laboratory criteria.^26,27^ Most patients with T1D at both centres use CSII with sensor augmentation or hybrid closed-loop systems, with CGM as standard practice also in children using a MDI regimen, consistent with ISPAD technology guidelines.^28,29^ Specialized neurophysiological evaluations include NCS and high-resolution nerve ultrasound, assessments that extend beyond routine clinical practice according to ISPAD guidelines.

### 4.2 Recruitment and Eligibility

The KIND cohort enrols individuals with T1D age 21 years and younger continuously at T1D diagnosis and during routine clinical visits. This creates a diverse cohort spanning the full spectrum of diabetes duration from newly diagnosed to long-standing disease in children and adolescents, enabling investigation of how the physiological developmental stage at T1D diagnosis influences diabetic peripheral neuropathy pathogenesis. Patients with other conditions affecting peripheral nerve function or structure are excluded to avoid influences other than T1D. These include premature birth (< 32 gestational weeks) or a family history of inherited neurological diseases.

The neurophysiology department at OKS routinely evaluates patients with unilateral fractures and suspected nerve trauma. As standard care, these patients undergo peripheral neuropathy screening (see 5.4) of the injured and uninjured extremity. The measurements taken from the uninjured side serve as the control for neurophysiological analyses.^9^ We applied this same screening protocol to our T1D cohort as described below.

### 4.3 Ethics and Consent

This study was approved by the Ethics Committee of Eastern Switzerland (approval number: 2022-00216, EKOS 22/018 and is registered with the Swiss project database. Ethics approval encompasses both prospective data collection and retrospective analysis of existing clinical data.

Age-appropriate informed consent procedures are followed, with written consent obtained from legal guardians for all patients younger than 18 years. Patients 18 years and older sign individual consent forms.

### 4.4 Follow-up Timeline

The KIND cohort is based on a longitudinal follow-up design that uses the established framework of clinical care at both participating centres (Figure 1), ensuing high retention rate, comprehensive data collection, and minimized patient burden.

**Figure 1.**
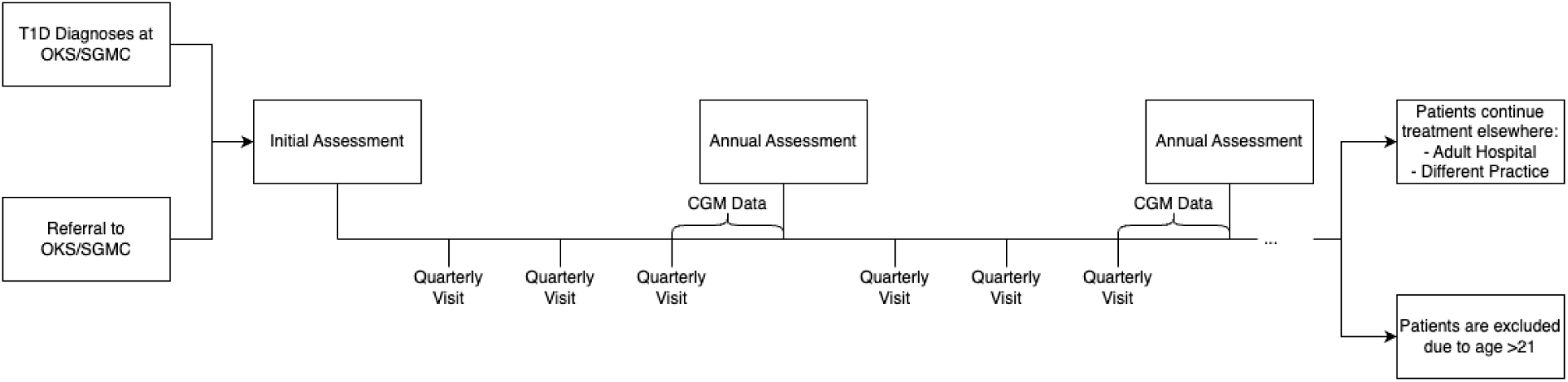
Overview of longitudinal study design.

**Figure 2.**
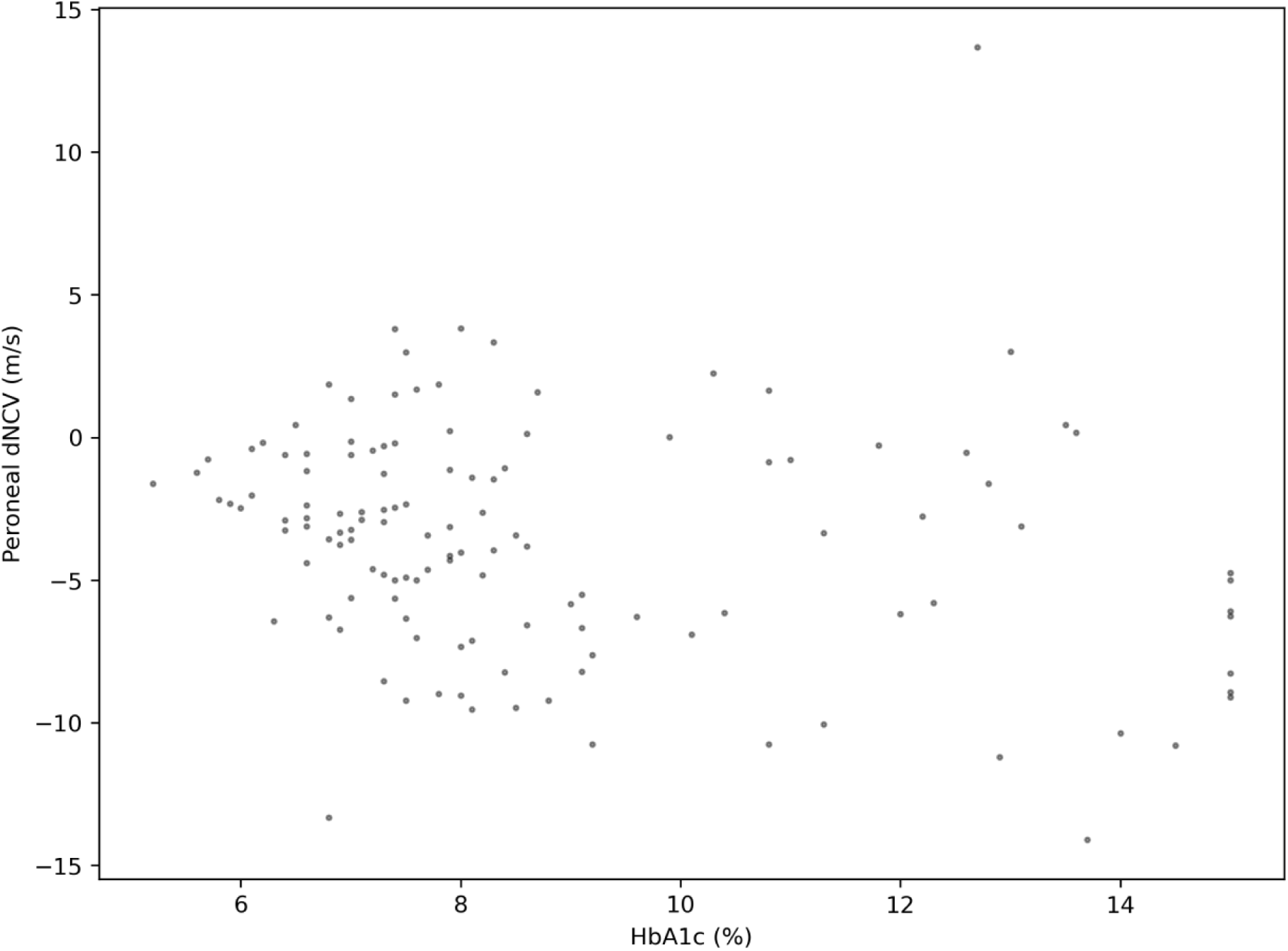
Peroneal dNCV and HbA1c at baseline

Following T1D diagnosis or initial enrolment, patients undergo a comprehensive baseline assessment that establishes detailed clinical, endocrinological, and specialized neurophysiological profiles including extensive laboratory testing. Subsequently, the study follows standard paediatric diabetes care protocols as outlined by ISPAD guidelines^30^ and previously described^9^, with quarterly visits as well as annual comprehensive assessments which include diabetes management assessment and the research-specific data collection.^18^ The anticipated primary reasons for study discontinuation are family relocation or transition to adult care.

## 5 DATA COLLECTION

Both participating centres utilize identical assessment protocols and comparable equipment, ensuring consistency in longitudinal measurements and enabling valid data pooling across sites. For neurophysiological assessments, patients registered at SGMC are referred to OKS, where specialized equipment and trained personnel are available to conduct NCS and high-resolution nerve ultrasound.

### 5.1 Clinical Measures

Standard growth parameters (height, weight, waist circumference) are measured using calibrated equipment following established protocols. These measurements enable calculation of derived indices including BMI, waist-to-height ratio, and body surface area (BSA; Du Bois formula^31^) expressed as sex- and age-adjusted variables (z-scores, referring to Swiss/WHO reference data^32^), if appropriate.

### 5.2 Diabetes Care

Assessments for diabetes care are presented in Table 1 and based on standardized protocols in accordance with ISPAD 2022 Clinical Practice Consensus Guidelines, incorporating 2024 updates.^33,34^ Glycaemic characterization combines quarterly HbA1c measurements with CGM data focused on 90-day windows aligned with the HbA1c integration period of two to three months^35^, enabling calculation and comparison of multiple GV metrics to determine their relative predictive value for neurophysiological outcomes. The comprehensive evaluation of the cohort characterizes diabetes management, growth patterns, and clinical status essential for investigating study aims.

**Table 1.** Data collection timeline and measurements *Note: CGM data collection occurs continuously, and data is exported and reviewed during quarterly visits

| Time Point | Quarterly Visits | Additional during Annual Assessment |
| --- | --- | --- |
| Clinical Measures | height, weight, blood pressure, heart rate | + waist circumference, pubertal stage |
| Diabetes Care | device/therapy review, total daily insulin dose, glucose monitoring and injection site examination | nothing additional to Quarterly Visits |
| CGM Data | data export and review* | nothing additional to Quarterly Visits |
| Laboratory | HbA1c, random capillary blood glucose | + random C-peptide, creatinine, lipid profile, electrolytes, screening for thyroid and celiac disease, albumin-creatinine ratio in urine. |
| Neurophysiology | - | + NCS, high-resolution nerve ultrasound, clinical assessment |

#### Physical Examination

Standardized examination includes blood pressure measurements, systematic assessment of insulin injection/infusion sites for lipohypertrophy, and annual clinical neuropathy screening using monofilament testing and vibration perception assessment.

#### Diabetes Management Assessment

Comprehensive documentation (Table 1) includes insulin delivery method (CSII versus MDI), device specifications (manufacturer, standalone/sensor-augmented/hybrid closed-loop systems), CGM device details (overview in **Error! Reference source not found.**), total daily insulin dose, insulin-to-carbohydrate ratios, and diabetes-related symptoms.

### 5.3 CGM Data Collection

#### Data Sources and Collection

CGM data is collected from systems commonly used in clinical practice: FreeStyle Libre 1/2/2+/3/3+ (Abbott Diabetes Care, Inc., Alameda, CA, USA), Dexcom G6/G7 (Dexcom, Inc., San Diego, CA, USA), and Medtronic Guardian 3/4/Simplera (Medtronic MiniMed™, Inc., Northridge, CA, USA). Data is accessed through manufacturer-specific platforms or the integrated diabetes management systems Glooko (Glooko AB, Gothenburg, Sweden), Tidepool (Tidepool Project, Palo Alto, CA, USA), and Diabass (mediaspects Beratungsgesellschaft für neue Medien GmbH, Balingen, Germany).

CGM data were collected as comprehensively as possible, with particular focus on the 90-day period preceding each neurophysiological assessment. This 90-day window aligns with the HbA1c integration period. When 90-day data is unavailable, shorter periods (14-90 days) can be used, given the high correlation between 14-day and long-term CGM metrics.^36^

Routine manual calibration of CGMs is not required unless specified by the device manufacturer (e.g. Medtronic) or indicated to parents/patients by suspected CGM inaccuracy.

#### Data Harmonization Challenge

Patients at OKS and SGMC use a variety of CGM devices (Table 3). Recent publications have highlighted performance differences and metric comparability issues across CGM manufacturers and devices,^20,37,38^ prompting standardized data pre-processing.

If necessary, CGM values are converted from mg/dl to mmol/L (conversion factor: 0.0555) and timestamps are standardized to Universal Time Coordinated before database storage.

Future analyses will require additional harmonization to address device-specific differences in measurement frequencies (ranging from 1 to 15-minute intervals) and out-of-range value handling (**Error! Reference source not found.**). Manufacturers use different measurement ranges and coding approaches. Definitions for temporal alignment and out-of-range values will be tailored to specific analytical objectives, as these decisions differentially impact glucose variability metrics. Additionally, statistical model specifications may include device manufacturer and model as a covariate to account for systematic measurement differences.

#### Glucose Metrics Calculation

Both clinically established (including mean glucose, standard deviation, coefficient of variation, time-in-range categories, time-in-tight-range) and advanced CGM metrics (including Continuous Overall Net Glycemic Action, Average Daily Risk Range, Glucose Fluctuation Moment Index)^17,19^ will be calculated according to previously published definitions (Supplementary Materials section **Error! Reference source not found. Error! Reference source not found.**).

### 5.4 Neurophysiology Assessment

#### 5.4.1 Nerve Conduction Studies

##### Assessment Protocol

Four standardized peripheral nerves are evaluated annually: peroneal, tibial, and median motor and median sensory nerves. The inclusion of motor and sensory components provides a comprehensive assessment, as sensory axons (which include small fibre components) outnumber motor axons by at least 9-to-1^39^ in peripheral nerves. This selection targets nerves with demonstrated sensitivity to diabetic neuropathy while ensuring reliable measurement across diverse paediatric developmental stages. The peroneal nerve receives particular attention given its pronounced vulnerability to early diabetic neuropathy.^9^

##### Standardized Protocol

All measurements follow established paediatric neurophysiology protocols with strict environmental standardization (room temperature-controlled at 21-23° C, measurement of skin temperature, standardized electrode placement, left extremity assessment unless contraindicated by injury or other factors requiring right-side assessment).40

Motor and sensory NCV and distal motor latency are the primary biomarkers. Secondary biomarkers include metrices for the dispersion of the compound muscle and sensory nerve action potential (CMAP & SNAP) and the total electric energy required to achieve the maximum CMAP amplitude.^41^

##### Paediatric-Specific Adaptations

To account for the established relationship between body height and NCV, we calculate height-adjusted difference values (*dNCV* = *measured NCV* − *expected NCV*; additional information in Appendix Chapter **Error! Reference source not found.**). Expected values are derived from adjustment models based on our healthy control group^9^, according to previously published models by Hyllienmark et al.^7,42,43^. This approach distinguishes diabetes-related impairment from developmental and environmental confounders. Negative dNCV values indicate performance below normative expectations and vice versa. A differentiated analysis on the relationship between peroneal NCV and height, age, and other influencing factors has been conducted.^9^ The relevance of temperature and similar analyses for the other three nerves are still ongoing.

#### 5.4.2 High-Resolution Ultrasound

##### Structural Assessment

High-resolution ultrasound provides direct visualization of nerve structure. High-frequency probes (Canon Aplio i800 with i22LH8 probes) are used to assess structural nerve biomarkers including cross-sectional area (CSA) and circumference of the median nerve.

##### Standardized Protocol

Median nerve ultrasound is performed at three standardized anatomical sites on the left arm (or right arm if contraindicated) in the fully supinated position: wrist (L1), mid-forearm (L2), and distal upper arm (L3).21,44,45 CSA at L2 serves as the primary outcome measure due to its sensitivity for early neuropathic changes.46

Transverse images are captured at each location and stored for offline analysis.^21^ Trained operators manually trace the nerve border (hyperechoic rim between nerve tissue and surrounding connective tissue) to measure CSA and circumference. This retrospective measurement approach minimizes patient discomfort and motion artifacts.

Additional metrics, including 3D median nerve reconstruction and median nerve tortuosity (analogous to corneal nerve assessments in diabetic neuropathy47,48), will be incorporated as methods become validated for ultrasound-based peripheral nerve assessment49.

##### Paediatric-Specific Adaptations

Median CSA is strongly correlated with body size50 and measurements are adjusted using body surface area calculations (using the Du Bois formula31) to account for normal growth-related changes in nerve size throughout childhood and adolescence51.

### 5.5 Laboratory Measurements

All laboratory measurements are taken during the annual assessment, except for HbA1c and random capillary blood glucose, which are measured quarterly as shown in Table 1.

HbA1c is measured using the Abbott AFINION™ 2 analyser, providing long-term data that complements CGM assessments. Random C-peptide is measured using chemiluminescence-immunoassay with the DiaSorin Liaison XL to assess residual beta-cell function, particularly valuable for capturing the heterogenous progression of beta-cell loss in patients with recently diagnosed T1D. Additionally, the capillary blood glucose levels are measured around the time of NCS (time difference ±1h).

Annual visits include comprehensive lipid profiles for metabolic risk assessment, thyroid and celiac disease for diabetic comorbidities screening, and the albumin-to-creatinine ratio in the first voided morning urine for diabetic complications (Table 1).

Electrolyte concentrations (sodium, potassium, chloride, magnesium and ionised calcium) are measured during the NCS appointment as important covariates for NCV.

## 6 PATIENT AND PUBLIC INVOLVEMENT

The study’s care-embedded design ensures research questions address clinical concerns from routine paediatric diabetes practice. The focus on glucose variability and early neuropathy detection arose from family concerns about CGM data interpretation and long-term complication risk expressed during clinic visits.

Families have been involved since study inception through presentations at public events and media engagement.52 We continuously collect feedback regarding participation burden and assessment preferences during routine clinical encounters.

Individual results are discussed with patients and families after each annual examination. Aggregate findings are shared annually at parent evenings and youth diabetes events, and through presentations at diabetes patient and parent association meetings and diabetes centre events.

## 7 PATIENT CHARACTERISTICS

Table 2 includes baseline demographic, anthropometric, and neurophysiological characteristics of the KIND cohort and the healthy control group. Table 3 provides diabetes specific measurements at baseline for the KIND cohort.

**Table 2.** Baseline characteristics of the control group and the KIND cohort All values are median and the first and third quartiles, unless otherwise specified; n for the number of patients; *overall percentage, not median; **preliminary results, see Appendix **Error! Reference source not found.**.

|  | Control Group<br>(n = 103) |  |  | KIND Cohort<br>(n=141) |  |  |
| --- | --- | --- | --- | --- | --- | --- |
|  | Median | [Q1; Q3] | n | Median | [Q1; Q3] | n |
| **Demographics & Anthropometrics** |  |  |  |  |  |  |
| Sex* |  |  |  |  |  |  |
| Female (%) | 53.4% |  | 55 | 47.5% |  | 67 |
| Male (%) | 46.6% |  | 48 | 52.5% |  | 74 |
| Age (year) | 10.9 | [7.5; 14.2] | 103 | 12.2 | [8.4; 14.3] | 141 |
| Height (cm) | 153.9 | [127.1; 162.1] | 66 | 152.8 | [131.2; 164.4] | 138 |
| Height (Z-Score) | 0.5 | [-0.1; 1.1] | 66 | 0.5 | [-0.1; 1.1] | 137 |
| Weight (kg) | 47.5 | [24.7; 57.6] | 65 | 42.1 | [26.8; 58.8] | 138 |
| Weight (Z-Score) | 0.3 | [-0.3; 1.3] | 64 | 0.3 | [-0.2; 1.0] | 136 |
| BMI (kg/m<sup>2</sup>) | 18.8 | [15.8; 22.2] | 65 | 18.2 | [16.0; 21.3] | 138 |
| BMI (Z-Score) | 0.3 | [-0.6; 1.2] | 65 | 0.4 | [-0.5; 1.0] | 137 |
| BSA (m<sup>2</sup>) | 1.4 | [0.9; 1.6] | 65 | 1.4 | [1.0; 1.6] | 138 |
| **NCS** |  |  |  |  |  |  |
| Temperature (°C) | 32.5 | [31.5; 33.0] | 33 | 31.8 | [30.9; 32.5] | 134 |
| NCV Peroneus (m/s) | 52.0 | [50.0; 55.0] | 54 | 48.0 | [46.0; 52.0] | 139 |
| dNCV Peroneus (m/s)** | -0.2 | [-1.9; 1.9] | 53 | -3.3 | [-6.2; -0.8] | 137 |
| NCV Tibialis (m/s) | 51.0 | [49.0; 53.0] | 45 | 48.0 | [45.0; 51.0] | 138 |
| dNCV Tibialis (m/s)** | -0.1 | [-1.8; 2.3] | 43 | -3.2 | [-6.5; -0.2] | 136 |
| NCV Medianus motor (m/s) | 58.0 | [55.2; 60.0] | 58 | 56.0 | [53.9; 58.0] | 140 |
| dNCV Medianus motor (m/s)** | 0.4 | [-2.8; 2.5] | 57 | -1.6 | [-4.0; 0.7] | 138 |
| NCV Medianus sensory (m/s) | 55.0 | [52.0; 61.0] | 52 | 54.0 | [50.0; 57.0] | 140 |
| dNCV Medianus sensory (m/s)** | -1.5 | [-3.6; 3.7] | 52 | -1.9 | [-6.6; 0.5] | 138 |
| **Ultrasound** |  |  |  |  |  |  |
| L1 Area (mm<sup>2</sup>) | 7.7 | [5.8; 10.0] | 78 | 5.9 | [4.8; 7.5] | 119 |
| L1 Circumference (mm) | 11.4 | [9.9; 12.8] | 75 | 11.2 | [9.9; 12.8] | 118 |
| L2 Area (mm<sup>2</sup>) | 5.8 | [4.5; 8.3] | 77 | 5.6 | [4.6; 6.8] | 125 |
| L2 Circumference (mm) | 9.0 | [8.1; 9.9] | 75 | 9.4 | [8.5; 10.4] | 125 |
| L3 Area (mm<sup>2</sup>) | 7.1 | [5.3; 8.5] | 36 | 7.0 | [5.4; 9.1] | 57 |
| L3 Circumference (mm) | 10.2 | [9.5; 11.9] | 34 | 11.2 | [9.8; 12.6] | 56 |

**Table 3.** Diabetes variables of the KIND cohort at baseline *CGM related data are for the 90 days preceding baseline NCS; **one patient has used CGM devices by two different manufacturers; ***reasons for “No CGM Data” include newly diagnosed patients, patients not wearing a CGM as part of their therapy, or no access to historical CGM data

|  | Median | [Q1; Q3] | n |
| --- | --- | --- | --- |
| **Basics** |  |  |  |
| Age at Diagnosis (year) | 7.9 | [4.3; 11.3] | 140 |
| Diabetes Duration (year) | 2.2 | [0.3; 5.1] | 140 |
| **Laboratory** |  |  |  |
| HbA1C (%) | 7.9 | [7.0; 9.9] | 132 |
| C-Peptide (pmol/L) | 17.0 | [0.0; 116.5] | 110 |
| Sodium (mmol/L) | 138.0 | [136.6; 139.0] | 58 |
| Potassium (mmol/L) | 4.1 | [4.0; 4.4] | 56 |
| Chloride (mmol/L) | 103.7 | [101.0; 105.4] | 52 |
| Ionized Calcium (mmol/L) | 1.2 | [1.2; 1.2] | 39 |
|  | Share |  | n |
| **CGM\*** |  |  |  |
| Patients with ≥ 70% CGM data available (%) | 40.4 |  | 57 |
| CGM Devices** |  |  |  |
| Abbott (%) | 27.0% |  | 38 |
| Dexcom (%) | 27.7% |  | 39 |
| Medtronic (%) | 18.4% |  | 26 |
| No CGM Data (%)*** | 27.7% |  | 39 |

## 8 FINDINGS TO DATE

Parts of this cohort have been previously published and presented at international conferences. As the cohort is continuously growing, patient numbers, characteristics, and inclusion/exclusion criteria may differ between analyses. Detailed information is available in the cited reports.

Cross-sectional NCS comparing 53 children with T1D (age 5.9-23.8 years) to 50 healthy controls (age 5.3-18.9 years) revealed significantly slower NCV across all four nerves examined (peroneal, tibial, median motor, and median sensory), with the peroneal nerve most severely affected.^9^ After adjusting for height, peroneal dNCV correlated most strongly with glucose SD (r=-0.45, p=0.0085), more so than with the coefficient of variation (r=-0.26, p=0.031) and HbA1c (r=-0.27, p=0.049). During the first 5 years post-diagnosis, dNCV declines as diabetes duration progresses (r=-0.41, p=0.0041) independently of glycaemic control, hypothesized to reflect the end of the remission phase and a decrease of C-peptide secretion.

Complementary structural assessment, namely measurement of the CSA of the median nerve in the middle of the forearm using high-resolution ultrasound, demonstrated median nerve enlargement at mid-forearm in children with T1D, particularly those with HbA1c >9%.46 After adjusting for body surface area (BSA), the mean CSA/BSA increased by 0.217 mm² per 1% HbA1c (p=0.004), with this association present even in children with diabetes duration <2 years, despite normal clinical examinations.

A previous longitudinal analysis of 45 children with serial NCS (median interval 17.9 [16.1; 25.8] months) demonstrated that changes in HbA1c (*HbA*1*c*_*t*1_ − *HbA*1*c*_*t*0_) were significantly and negatively associated with changes in peroneal dNCV (*dNCV*_*t*1_ − *HdNCV*_*t*0_; β=-0.59, p=0.014). In most patients, dNCV decreased with increasing HbA1c. Other patients showed improvements in dNCV with decreasing HbA1c, suggesting potential reversibility of early functional nerve impairment with improved glycaemic control.^19^

Figure 3 shows the evolution of peroneal dNCV across diabetes duration. Some patients exhibit a successive degradation of peroneal dNCV while others appear to improve over time. There are also multiple instances of an initial drop followed by a rebound in dNCV. One female patient, as an example, is exhibiting normal to very high dNCV across all four nerves (peroneal=13.67 m/s; tibial=-0.14m/s; median motor=0.25; median sensor=8.36m/s) despite high HbA1c measured six days after diagnosis (Age=14.2 y; HbA1c=12.7 %; C-Peptide=374 pmol/L; pH=7.38). These observations raise further questions, and we hope to be able to provide new insights in the course of this cohort study.

**Figure 3.**
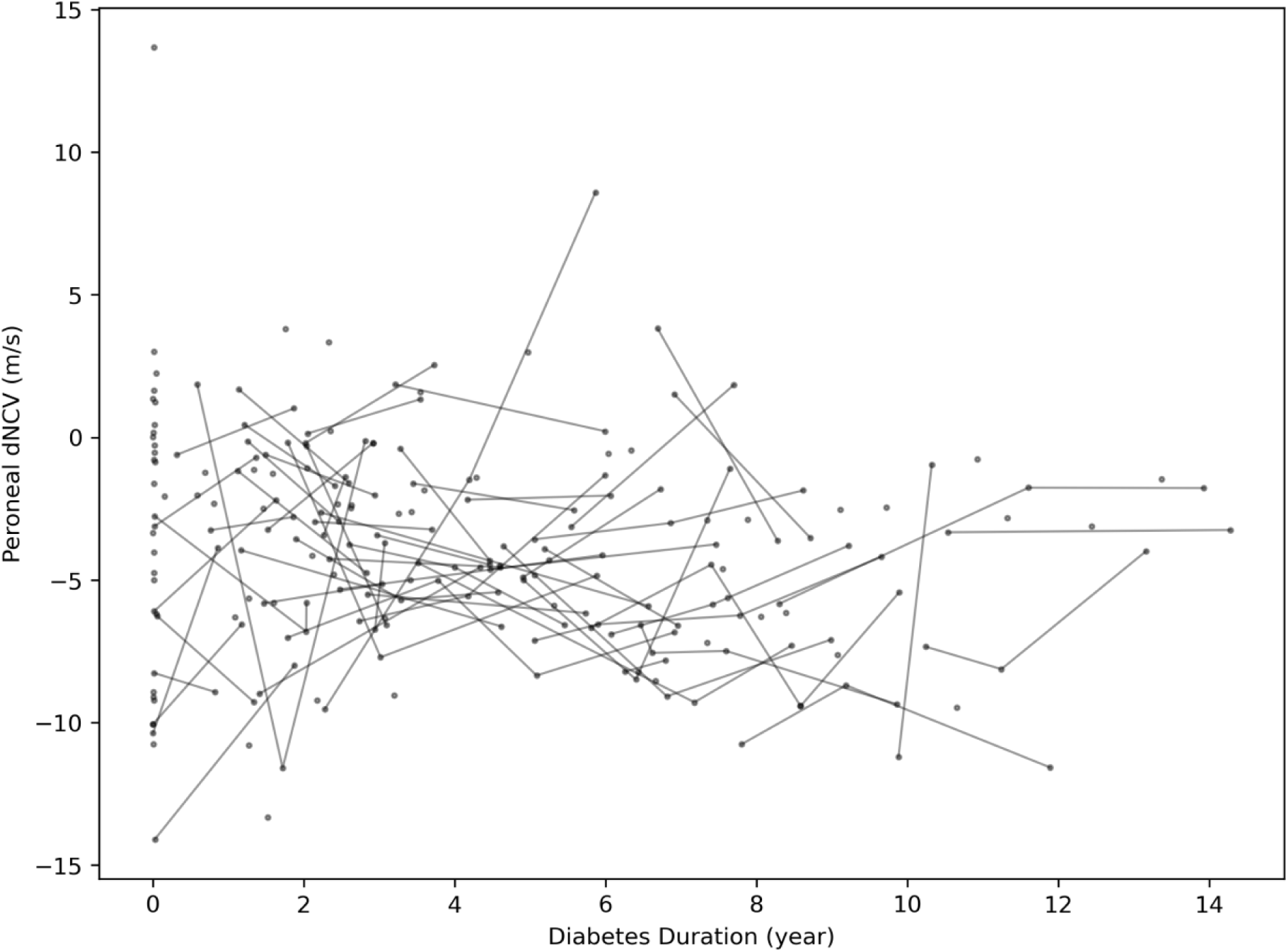
Peroneal dNCV across diabetes duration Connected dots represent observations for the same patient

To date, 141 T1D patients have completed at least one, 64 at least two, 24 at least three, five at least four (over 3.5 - 5.1 years), and one patient has completed five assessments (over 3.7 years).

## 9 STRENGTHS AND LIMITATIONS

### Strengths

The care-embedded longitudinal design of the KIND cohort with continuous enrolment from T1D diagnosis minimizes participant burden, maximizes retention, and captures neuropathy from its earliest clinically unapparent stages through adolescence, covering the full spectrum of diabetes duration and developmental stages. The study employs comprehensive non-invasive neurophysiological assessment combining functional (NCS across four nerves) and structural (high-resolution ultrasound) measures, with development-adjusted analyses (dNCV, BSA-adjusted CSA) that isolate diabetes-related impairment from normal growth using healthy control reference values. The longitudinal design with detailed glycaemic characterization through quarterly HbA1c and continuous CGM data access enables direct comparison of established and advanced glycaemic variability metrics (including GFMI and other complex indices) as predictors of neurophysiological outcomes over time, complemented by C-peptide assessment to investigate residual beta-cell function effects.

### Limitations

This regional two-centre study has limited geographic diversity that may affect generalizability. While the sample size is substantial for paediatric diabetic neuropathy research, the heterogeneous nature of the paediatric population (wide age range, developmental stages, and diabetes duration) combined with multiple influencing factors limits statistical power for various analyses. CGM device heterogeneity across multiple manufacturers and models with different measurement characteristics complicates metric standardization and comparability. Additionally, the control cohort comprises a convenience sample of unilateral fracture patients rather than matched healthy volunteers.

## 10 FURTHER DETAILS

### 10.1 Collaboration

An extension of the KIND cohort is planned in collaboration with Prof. Dr. Felix Wortmann and Prof. Dr. Tobias Kowatsch from the University of St. Gallen starting in mid 2026, integrating physical activity measures, supported by the Swiss National Science Foundation (Grant Number: 10006264).

We welcome future collaboration with other researcher teams. Interested researchers can reach out to the corresponding author for additional information.

### 10.2 Data Sharing Statement

Data are available upon reasonable request and in compliance with national, institutional, ethical, and informed consent agreements given the vulnerability of the cohort.

### 10.4 Author Contributions

Marc Robin Gruener: Conceptualization, Data Curation, Formal Analysis, Funding Acquisition, Investigation, Methodology, Project Administration, Software, Validation, Visualization, Writing (Original Draft Preparation)

Erin West: Conventionalization, Data Curation, Formal analysis, Methodology, Validation, Visualization

Ute Muhitira: Data Curation, Investigation, Methodology, Resources, Writing (Review & Editing)

Katrin Heldt: Data Curation, Investigation, Methodology, Resources, Writing (Review & Editing)

Sarah Oberhauser: Data Curation, Investigation, Methodology, Resources

Dagmar l’Allemand: Conceptualization, Funding Acquisition, Methodology, Resources, Supervision, Writing (Original Draft Preparation + Review & Editing)

Philip Broser: Conceptualization, Data Curation, Formal Analysis, Funding Acquisition, Investigation, Methodology, Project Administration, Resources, Supervision, Validation, Writing (Review & Editing)

## Acknowledgements

We thank all participating children and families for their essential contributions to this cohort. We are grateful to the entire KIND study group for their support during the data collection and clinical implementation.

