## Supplement for "The KIND cohort profile: longitudinal assessment of glycaemic management and neurophysiological outcomes in paediatric type 1 diabetes in Switzerland"

### 1 SUPPLEMENTARY MATERIALS

#### 1.1 Height adjusted nerve conduction velocity (dNCV)

As outlined in **Error! Reference source not found.** and previously described in Oberhauser et al.[9], we define *height adjusted Nerve Conduction Velocity* (*dNCV)* as the difference between the expected NCV for a child of a certain height $h$ in cm and the measured NCV ($dNCV = NCV(h)_{expected} - NCV_{measured}$). The expected NCV is determined by fitting a linear regression model $NCV_{expected} \sim Height\ (cm)$ to the data obtained from our healthy control group, adjusted according to Hyllienmark et al.[7,42,43]. The relevance of age and other influencing factors influencing factors (e.g. temperature) in addition to height is currently being evaluated for the peroneal, tibial as well as median motor and sensory nerves in the T1D cohort.

*Supplementary Table 1: Linear regression models for the healthy control group*

| Nerve | Intercept | β Height | P>\|t\| | 95% CI | n |
|---|---|---|---|---|---|
| Peroneus (m/s) | 62.2675 | -0.0671 | 0.001 | [-0.106; -0.028] | 53 |
| Tibialis (m/s) | 51.8021 | -0.0040 | 0.899 | [-0.068; 0.060] | 43 |
| Medianus motor (m/s) | 46.5655 | 0.0748 | 0.002 | [0.028; 0.122] | 58 |
| Medianus sensory (m/s) | 45.0256 | 0.0774 | 0.017 | [0.015; 0.140] | 50 |

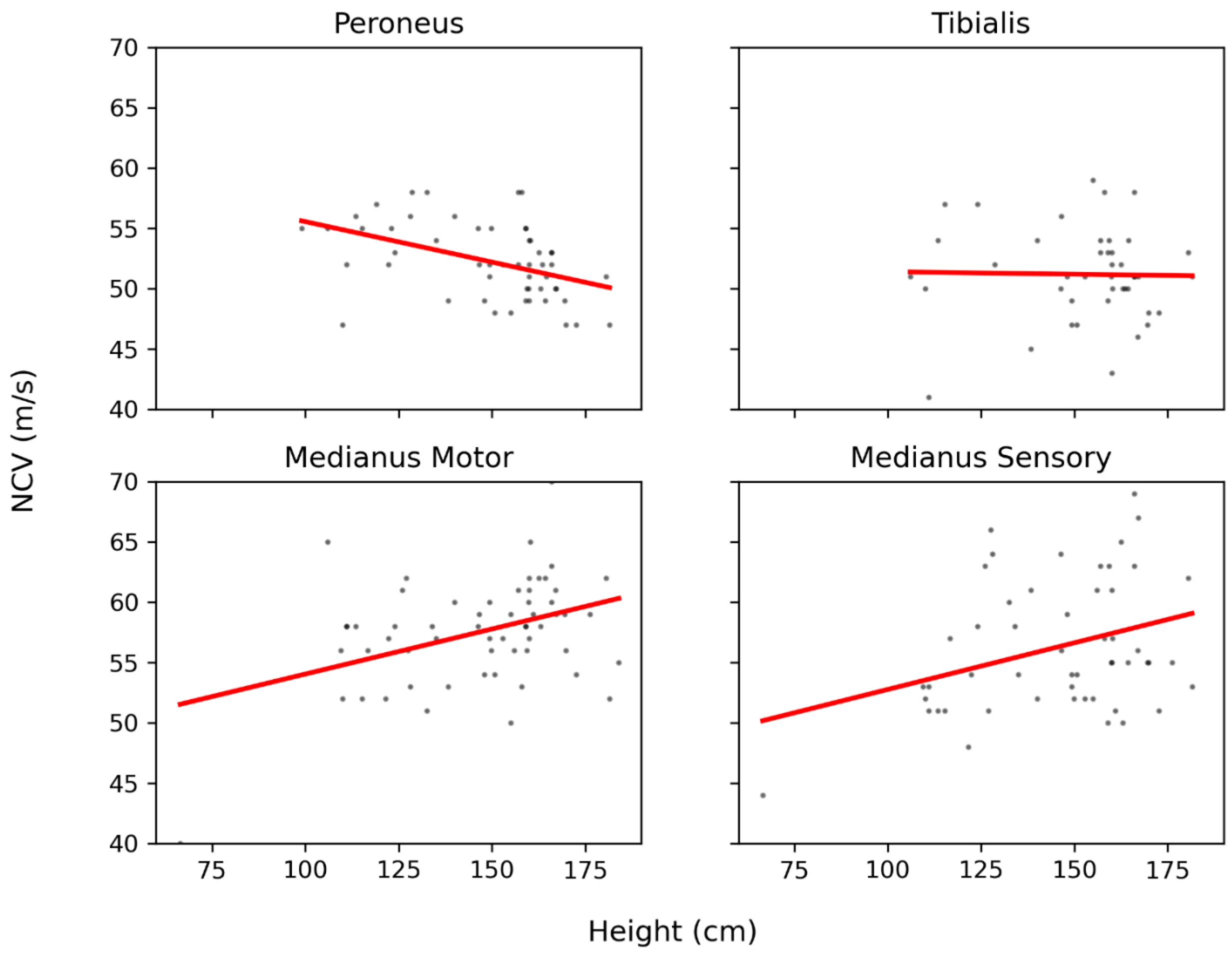


*Supplementary Figure 1: NCV compared to height in healthy controls*

#### 1.2 Comparison of different CGM Sensors

As mentioned above, there are relevant differences in the characteristics of different CGM devices and manufacturers (Supplementary Table 2) and derived metrics[20].

*Supplementary Table 2: Comparison of different CGM sensors*
*(MARD = Mean Absolute Relative Difference)*

#### CGM Comparison

| Manufacturer | Device | Wear Time (days) | Warm-up Duration (h) | Calibration | Manual Calibration possible | Official Measurement Range | Measurement Period | Age | MARD | | Source |
|---|---|---|---|---|---|---|---|---|---|---|---|
| | | | | | | | | | Adult | Children & Adulescent | |
| Abbott | FreeStyle Libre 2 FGM | 14 | 1 | factory-calibration | ❌ | 2.2 - 22.2 mmol/L | 1 min | > 4 | 8.10 % | 8.20 % | https://www.accessdata.fda.gov/cdrh_docs/reviews/K233537.pdf<br>https://www.accessdata.fda.gov/cdrh_docs/pdf22/K223435.pdf |
| Abbott | FreeStyle Libre 2+ FGM | 15 | 1 | factory-calibration | ❌ | 2.2 - 22.2 mmol/L | 1 min | > 2 | | | https://www.accessdata.fda.gov/cdrh_docs/reviews/K233537.pdf<br>https://www.accessdata.fda.gov/cdrh_docs/pdf22/K223435.pdf |
| Abbott | FreeStyle Libre 3 CGM | 14 | 1 | factory-calibration | ❌ | 2.2 - 22.2 mmol/L | 1 min | > 4 | 6.80 % | 7.80 % | https://www.accessdata.fda.gov/cdrh_docs/reviews/K212132.pdf<br>https://www.accessdata.fda.gov/cdrh_docs/pdf22/K223435.pdf |
| Abbott | FreeStyle Libre 3+ CGM | 15 | 1 | factory-calibration | ❌ | 2.2 - 22.2 mmol/L | 1 min | > 2 | | | https://www.accessdata.fda.gov/cdrh_docs/reviews/K212132.pdf<br>https://www.accessdata.fda.gov/cdrh_docs/pdf22/K223435.pdf |
| Dexcom | Dexcom G6 | 10 | 2 | calibration-code | ✅ | 2.2 - 22.2 mmol/L | 5 min | > 2 | 8.90% | 10.70 % | https://www.accessdata.fda.gov/cdrh_docs/reviews/DEN170088.pdf |
| Dexcom | Dexcom G7 | 10 | 0.5 | calibration-code | ✅ | 2.2 - 22.2 mmol/L | 5 min | > 2 | 8.90% | 9.30 % | https://www.accessdata.fda.gov/cdrh_docs/reviews/K213919.pdf |
| Medtronic | Guardian 3 | 7 | 2** | manual-calibration | ✅ | 2.2 - 22.2 mmol/L | 5 min | CH: > 2<br>US: > 14 | 10.55 %*** | | https://www.accessdata.fda.gov/cdrh_docs/pdf16/P160017S017B.pdf<br>https://www.accessdata.fda.gov/cdrh_docs/pdf16/P160017S031C.pdf |
| Medtronic | Guardian 4 | 7 | 2 | factory-calibration | ✅ | 2.8 - 22.2 mmol/L | 5 min | CH: > 2<br>US: > 7 | 10.60 % | 11.60 % | https://www.accessdata.fda.gov/cdrh_docs/pdf16/P160017S091B.pdf<br>https://www.accessdata.fda.gov/cdrh_docs/pdf16/P160017S091C.pdf |
| Medtronic | Simplera | 6+1* | 2 | factory-calibration | ✅ | 2.8 - 22.2 mmol/L | 5 min | CH: > 2<br>US: > 18 | 10.20 % | | https://www.accessdata.fda.gov/cdrh_docs/pdf16/P160007S047c.pdf<br>https://www.accessdata.fda.gov/cdrh_docs/pdf16/P160007S047b.pdf<br>https://eud.sukl.gov.cz/pub/deska/40000001/athena/25V0026H%40SUKLAA/0001011954%40ISZPBP2/Simplera%20Sensor%20MDR_DoC.pdf<br>https://www.diabetesschweiz.ch/fileadmin/user_upload/01_Betroffene_und_Angehoerige/Services/Technische_Hilfsmittel/CGM_Systeme_DE_2025.pdf |

*24h Grace Period ; ** 5 min after first calibration request which should be done 2h post-insertion; *** Study included adolescents and adults aged 15-75

#### 1.3 Advanced Glucose Variability Metrics

The following glucose variability metrics are briefly described in terms of their potential ability to predict slowing of NCV as early sign of subclinical diabetic peripheral neuropathy in children and adolescents with T1D.

The Continuous Overall Net Glycemic Action (CONGAn) measures intraday glycaemic variation through glucose differences at defined time intervals. Validated in paediatric Type 1 diabetes populations, this metric enables flexible examination of both short- and long-term glucose patterns.[16]

The Average Daily Risk Range (ADRR) accounts for asymmetric glucose variations through distinct risk functions for hypoglycaemia and hyperglycaemia. Logarithmic transformation enables balanced evaluation of extreme glycaemic events and overall risk.[53–55]

The Mean Absolute Glucose Change (MAG) captures the total magnitude of glucose fluctuations over time, distinguishing between profiles with comparable means or standard deviations but different control patterns. This metric proves particularly useful for detecting clinically relevant variations in intensive care environments.[56]

The Glycemic Variability Percentage (GVP) employs a geometric approach, comparing actual glucose trajectory lengths to theoretical minimum lengths. This method specifically identifies high-frequency and high-amplitude fluctuations, offering complementary insight into glucose stability.[57]

We introduce the Glucose Fluctuation Moment Index (GFMI), combining squared glucose change velocity with squared deviation from the 5 mmol/L target.[19] The GFMI highlights substantial glucose shifts occurring distant from target range:

$$GFMI = \frac{1}{n}\sum_{i=1}^{n}(g_{t=i+1} - g_{t=i})^2 * (g_{t=i} - 5)^2$$

*Supplementary Table 3: Characteristics of selected advanced glucose variability metrics (Reproduction of Gruener et al.[58])*

| Metric | Definition and Purpose | Formula | Key Clinical & Scientific Applications | Distinguishing Features |
|---|---|---|---|---|
| Continuous Overall Net Glycemic Action (CONGAn)[16] | Measures glycaemic variation by analysing differences between glucose readings at specified time intervals (n hours) | $CONGAn = \sqrt{\frac{\sum_{t=t_1}^{t_{k^*}}(D_t - \overline{D})^2}{k^* - 1}}$<br>$with\ D_t = GR_t - GR_{t-m}$<br>$and\ \overline{D} = \frac{\sum_{t=t_1}^{t_{k^*}} D_t}{k^*}$<br>$and\ k^* = n\ x\ 60min$ | Association with distal symmetric polyneuropathy in T1D children; Validation of synthetic CGM data; Assessment of physical activity impact on glycaemic variability [59–64] | Flexible time intervals (1h, 2h, 4h, 12h, 24h) for different clinical objectives; captures temporal glucose dynamics |
| Average Daily Risk Range (ADRR)[53–55] | Quantifies combined risk of hypo- and hyperglycaemia through logarithmic transformation of glucose values | $ADRR = \frac{1}{M}\sum_{i=1}^{M}[LR^i + HR^i]$<br>$with\ LR^i = max[rl(x_1^i),\ldots,rl(x_n^i)]\ and\ HR^i = max[rh(x_1^i),\ldots,rh(x_n^i)]\ for\ day\ i; = 1,2,\ldots,M$<br>$and\ rl(BG) = r(BG) if\ f(BG) < 0\ else\ 0$<br>$and\ rh(BG) = r(BG)\ if\ f(BG) > 0\ else\ 0$<br>$and\ r(BG) = f(BG) = 1.509 * [ln(BG)^{1.084} - 5.381]$ | Evaluation of T1D and T2D treatment regimens; Mortality prediction in burn ICU patients; Assessment of diabetes support systems[59,60,65–68] | Addresses skewness in CGM data through logarithmic transformation; combines both high and low glucose risks |
| Mean Absolute Glucose Change (MAG)[56] | Measures the sum of absolute glucose changes over time, designed to capture clinically relevant glucose variations | $MAG = \frac{\|\Delta BG\|}{\Delta Time}$ | Prediction of ICU mortality; Assessment of critically ill non-diabetic children; Classification of gastroparesis in T1D[56,69,70] | Differentiates between clinically distinct glucose patterns that may have similar means/SDs; Particularly relevant for ICU settings |

| | | | | |
|---|---|---|---|---|
| Glycemic Variability Percentage (GVP)[57] | Compares actual glucose trace length to minimal possible length, emphasizing oscillating excursions | $GVP = (\frac{L}{L_0} - 1) * 100$ $with\ L = \sum_{i=1}^{n} \sqrt{\Delta x_i^2 + \Delta y_i^2}$ $and\ L_0 = \sum_{i=1}^{n} \Delta x_i$ | Differentiation between T2D and impaired glucose tolerance; Prediction of nocturnal hypoglycaemia; Evaluation of automated insulin delivery[71–74] | Penalizes high-frequency and high-amplitude oscillations; Geometric approach to variability assessment |
| Glucose Fluctuation Moment Index (GFMI)[75] | Integrates velocity of glucose change weighted by distance from glycaemic target | $GFMI = \frac{1}{n} \sum_{i=1}^{n} (g_{t=i+1} - g_{t=i})^2 * (g_{t=i} - 5)^2$ | Under investigation for association with nerve vitality[75] | Uniquely weights changes by distance from target; Emphasizes substantial changes in glucose concentration away from target range |

#### 1.4 Subgroup CGM characteristics of the KIND Cohort

Data was filtered to only include the first observation for KIND cohort participants with ≥70% CGM data available for the 90-days preceding the NCS appointment. Data was resampled from 1, 5, or 15min, respectively, to 15min periods (via forward-fill with a limit of 1) where 100% data availability equates to 8’640 CGM ($90\ days\ *\ 24\ h/day\ *\ 60\ min/h\ /\ 15min\ =\ 8'640$) values post-resampling. Values out of measurement range were marked as missing. After filtering, 81 participants remained.

*Supplementary Table 4: Subgroup characteristics of 81 participants at the first observation for with ≥70% CGM data availability*
**100% equals 8’640 CGM values after resampling to 15 min*

| | **Median** | **[Q1; Q3]** |
|---|---|---|
| Data Availability (%)* | 95.45 | [86.98; 97.99] |
| CGM values excluded “low” (%)* | 0.07 | [0.00; 0.38] |
| CGM values excluded “high” (%)* | 0.91 | [0.21; 5.67] |
| Mean Glucose (mmol/L) | 9.05 | [8.51; 9.77] |
| Median Glucose (mmol/L) | 8.40 | [7.80; 9.20] |
| Glucose Standard Deviation | 3.67 | [3.17; 4.15] |
| Glucose CV (%) | 38.73 | [36.64; 42.52] |
| TIR | | |
| very low (%) | 0.41 | [0.18; 0.75] |
| low (%) | 2.11 | [1.12; 3.52] |
| normal (%) | 61.16 | [54.17; 70.94] |
| high (%) | 22.26 | [20.22; 25.90] |
| very high (%) | 11.02 | [6.11; 18.10] |
| ADRR | 45.08 | [38.04; 53.34] |
| CONGA24 | 4.56 | [4.06; 5.34] |
| GVP | 29.07 | [19.20; 33.74] |
| MAG | 3.20 | [2.64; 3.66] |
| GFMI | 45.29 | [30.15; 75.48] |
